# Magnitude and time-course of excess mortality during COVID-19 outbreak: population-based empirical evidence from highly impacted provinces in northern Italy

**DOI:** 10.1101/2020.07.10.20150565

**Authors:** Sara Conti, Pietro Ferrara, Giampiero Mazzaglia, Marco I. D’Orso, Roberta Ciampichini, Carla Fornari, Fabiana Madotto, Michele Magoni, Giuseppe Sampietro, Andrea Silenzi, Claudio V. Sileo, Alberto Zucchi, Giancarlo Cesana, Lamberto Manzoli, Lorenzo G. Mantovani

## Abstract

**Background:** The real impact of SARS-CoV-2 on overall mortality remains uncertain and surveillance reports attributed to COVID-19 a limited amount of deaths during the outbreak. Aim of this study is to assess the excess mortality (EM) during COVID-19 outbreak in highly impacted areas of northern Italy.

**Methods:** We analyzed data on deaths occurred in the first four months of 2020 in health protection agencies (HPA) of Bergamo and Brescia (Lombardy), building a time-series of daily number of deaths and predicting the daily standardized mortality ratio (SMR) and cumulative number of excess deaths (ED) through a Poisson generalized additive model of the observed counts in 2020, using 2019 data as a reference.

**Results:** We estimated 5740 (95% Credible Set (CS): 5552–5936) ED in the HPA of Bergamo and 3703 (95% CS: 3535 – 3877) in Brescia, corresponding to 2.55 (95% CS: 2.50–2.61) and 1.93 (95% CS: 1.89–1.98) folds increase in the number of deaths. The ED wave started a few days later in Brescia, but the daily estimated SMR peaked at the end of March in both HPAs, roughly two weeks after the introduction of lock-down measures, with significantly higher estimates in Bergamo (9.4, 95% CI: 9.1–9.7).

**Conclusion:** EM was significantly larger than that officially attributed to COVID-19, disclosing its hidden burden likely due to indirect effects on health system. Time-series analyses highlighted the impact of lockdown restrictions, with a lower EM in the HPA where there was a smaller delay between the epidemic outbreak and their enforcement.

## Introduction

As the novel coronavirus disease 2019 (COVID-19) continues to spread internationally since the beginning of the outbreak in December 2019, one of the most relevant public health concerns has been its mortality rate [1,2].

Italy is among the countries with the highest number of severe acute respiratory syndrome coronavirus 2 (SARS-CoV-2) infections, reaching more than 240,000 confirmed cases and around 35,000 deaths in the late June [3]. According to the epidemiological bulletin of June 25^th^ of the Italian National Institute of Health, patients who died of COVID-19 had a median age of 82 years, while the median age of subjects who were positive to SARS-CoV-2 infection was 62 years [4,5]. Surveillance data also suggested that principal risk factors for mortality in COVID-19 patients were older age, male sex, and concomitant conditions [4,6-8].

Current estimates of case fatality ratio (CFR) diverge considerably from one country to another, likely due to differences in age distribution and health status of the population. There is also general consensus that CFR variability may be explained by inaccurate estimates in the number of people who are infected with the SARS-CoV-2. Asymptomatic cases of COVID-19, patients with mild symptoms, or individuals who are misdiagnosed could be left out of the denominator, leading to its underestimation and overestimation of the CFR [1,9]. Thus, at the moment CFRs do not provide reliable assumption to assess the real impact of COVID-19 on country-based mortality burden [10].

Moreover, early research assessed that the mortality burden during the first phase of the COVID-19 spread in Italy is underestimated, because epidemiological surveillance reports have been attributing to COVID-19 only a limited amount of all deaths [1,2,13,14], suggesting that SARS-CoV-2 epidemic had a bigger impact on overall mortality by direct and indirect effects [1,2]. The former may encompass an important proportion of people who would eventually die from the disease before the diagnosis was made; while the latter, indirect effects, must be mainly sought within the patient comorbid status, as well as the disproportionate hospital overload and high shortage of healthcare resources in the most affected areas of the country [2,13].

In particular, analyses on mortality data published by the Italian Institute of Statistics (ISTAT) found increased standardized rate ratios of mortality across the different areas of the country within the first weeks of the COVID-19 epidemic. This excess mortality was found to be majorly diffused in the northern Italian regions [2]. Here, Lombardy registered the biggest COVID-19 outbreak in Italy (and thus in Europe), accounting for more than 94,000 diseases cases [3], and some of its provinces – including Bergamo and Brescia – have been the hardest-hit for the death toll during the peak epidemic weeks [13,14].

Scarce evidence is available on excess overall mortality during the outbreak using population-based direct empirical observations and little or nothing has been published on the time-course of this excess.

Based on these considerations, we therefore conducted a retrospective analysis of mortality data of a densely populated vast geographical area of northern Italy in the first four months of 2020, with the aim to describe the magnitude and time-course of entire cycle of the excess mortality associated with COVID-19 outbreak, also investigating the role of lockdown measures in the mitigation of the disease impact on the general mortality.

## Methods

### Study population

Lombardy region, the epicenter of the Italian SARS-CoV-2 outbreak, is served by eight local health protection agencies (HPA), which cover mutually exclusive areas of the region.

We focused on all inhabitants of the HPAs of Bergamo and Brescia, where the outbreak magnitude has been particularly intense [13,14].

### Study design and data sources

Italy has a tax-based, universal coverage National Health System organized in three levels: National, Regional, and Local. In Lombardy, the local HPAs are primarily responsible to collect healthcare information (e.g. inpatient and outpatient treatments), including mortality data of all inhabitants under their jurisdiction [15].

We carried out a longitudinal retrospective time-series study on the overall mortality traced in these standardized healthcare regional administrative databases.

To accomplish the study objectives, we retrieved data on all-cause deaths occurred between January 1^st^ and April 30^th^ in the years 2020 and 2019, the latter to be used as a reference period. For each deceased subject we linked the unique identification code in the mortality database with demographic information (i.e. age at death and gender).

We chose 2019 as a reference, because we observed that the seasonality of the flu outbreak, that is one of the driver of variations in mortality during winter, was similar to that observed in 2020 up to the beginning of COVID outbreak [16], while during previous years peaks occurred in different periods. Therefore, we hypothesize that 2019 fairly approximates the expected situation in 2020.

### Statistical analysis

We compared the age and sex distribution among deceased in 2019 and 2020 through chi-square tests, and the number of daily deaths through Wilcoxon rank-sum test.

We built the time-series of the number of observed daily deaths during January-April 2019 and 2020 for each HPA. Based on these time-series, and considering that the population of each HPA remained stable from 2019 to 2020 (HPA Brescia: from 1,089,602 to 1,097,648; HPA Bergamo: from 1,160,374 to 1,163,243), we computed the expected daily number of deaths for 2020 as the average daily number of deaths occurred in 2019.

In order to assess the magnitude and the time course of the mortality variation during SARS-CoV-2 outbreak, we built a Poisson generalized additive model, with the daily counts of deaths during 2020 as the outcome, and the HPA, a non-linear function of date and the number of expected deaths as the predictors.

In detail, the model was:

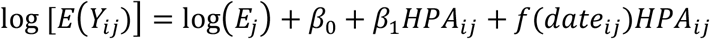

where *Y*_*ij*_ is the observed number of deaths during the *i*-th day in *j*-th HPA, *E*_*j*_ is the daily expected number of deaths in the *j*-th HPA, and it is held fixed for each *i*-th day, *HPA*_*ij*_ is a categorical variable indicating the HPA where the number of deaths was measured in the *i*-th day, and *f*(*date*_*ij*_) is a thin plate regression spline of the calendar date [17]. The term *f*(*date*_*ij*_)*HPA*_*ij*_ indicates that a separate spline is fitted for each HPA, allowing for the estimation of a different epidemic time course in the two areas. The use of the constant *E*_*j*_ as an offset allows to directly model the daily Observed/Expected ratio of mortality, that is the Standardized Mortality Ratio (SMR), instead of the daily counts, predicting both the estimated number of daily deaths and the estimated SMR [18,19].

Therefore, based on the model results, we first predicted the SMR for each day of the 2020, together with its 95% Confidence Interval (CI). Thereafter, using the method reported by Rivera et al. [20], we estimated the cumulative number of excess deaths, as compared to 2019, and their correlated 95% Credible Sets (CS), which similarly to CI synthesize the precision of the estimates but based on Bayesian methods. Finally, we expressed the excess in term of n-fold increase in the cumulative number of deaths, using 2019 cumulative number of deaths as the reference.

We replicated the whole analysis stratifying by sex and by age, categorized as: < 60 years, 60-69 years, 70-79 years, 80-89 years, and 90+ years.

All analyses were carried out with statistical software SAS version 9.4 (SAS Institute, Cary, NC, USA) and R version 4.0.0 (R Project for Statistical Computing, www.R-project.org).

## Results

During the first four months of 2020, 17,099 deaths were registered among the residents of the two HPAs of Bergamo and Brescia, as compared to 7592 in 2019 (**Table 1**). In both provinces, the crude number of deaths increased by more than 90%. Overall, the daily average number of deaths significantly rose from 63 (SD: 10.7) to 141 (SD: 129.8), with a steeper wave of the cumulative time-series analysis in the area of Bergamo (**Figure 1**).

**Table 1.** **Characteristics of the deceased over the period January 1^st^ - April 30^th^, stratified by Health Protection Agency (HPA). Comparison between 2020 and 2019.**

|  | HPA Bergamo |  | HPA Brescia |  | Total |  |
| --- | --- | --- | --- | --- | --- | --- |
|  | 2019 | 2020 | 2019 | 2020 | 2019 | 2020 |
| **N deaths** | 3662 | 9433 | 3930 | 7666 | 7592 | 17099 |
| **Gender** |  |  |  |  |  |  |
| Female | 1920 (52.4) | 4407 (46.7) | 2082 (53.0) | 3823 (49.9) | 4002 (52.7) | 8230 (48.1) |
| Male | 1742 (47.6) | 5026 (53.3) | 1848 (47.0) | 3843 (50.1) | 3590 (47.3) | 8869 (51.9) |
| *p-value vs 2019* |  | <0.001 |  | 0.002 |  | <0.001 |
| **Age class** |  |  |  |  |  |  |
| <60 | 281 (7.7) | 440 (4.7) | 242 (6.2) | 357 (4.7) | 523 (6.9) | 797 (4.7) |
| 60-69 | 321 (8.8) | 873 (9.3) | 313 (8.0) | 634 (8.3) | 634 (8.4) | 1507 (8.8) |
| 70-79 | 741 (20.2) | 2267 (24.0) | 786 (20.0) | 1713 (22.3) | 1527 (20.1) | 3980 (23.3) |
| 80-89 | 1399 (38.2) | 3809 (40.4) | 1543 (39.3) | 3019 (39.4) | 2942 (38.8) | 6828 (39.9) |
| 90+ | 920 (25.1) | 2044 (21.7) | 1046 (26.6) | 1943 (25.3) | 1966 (25.9) | 3987 (23.3) |
| *p-value vs 2019* |  | <0.001 |  | <0.001 |  | <0.001 |
| **Daily number of deaths** |  |  |  |  |  |  |
| Mean (SD) | 30.5 (6.39) | 78.0 (82.75) | 32.8 (6.94) | 63.4 (49.49) | 63.3 (10.65) | 141.3 (129.80) |
| *p-value vs 2019* |  | <0.001 |  | <0.001 |  | <0.001 |

**Figure 1.**
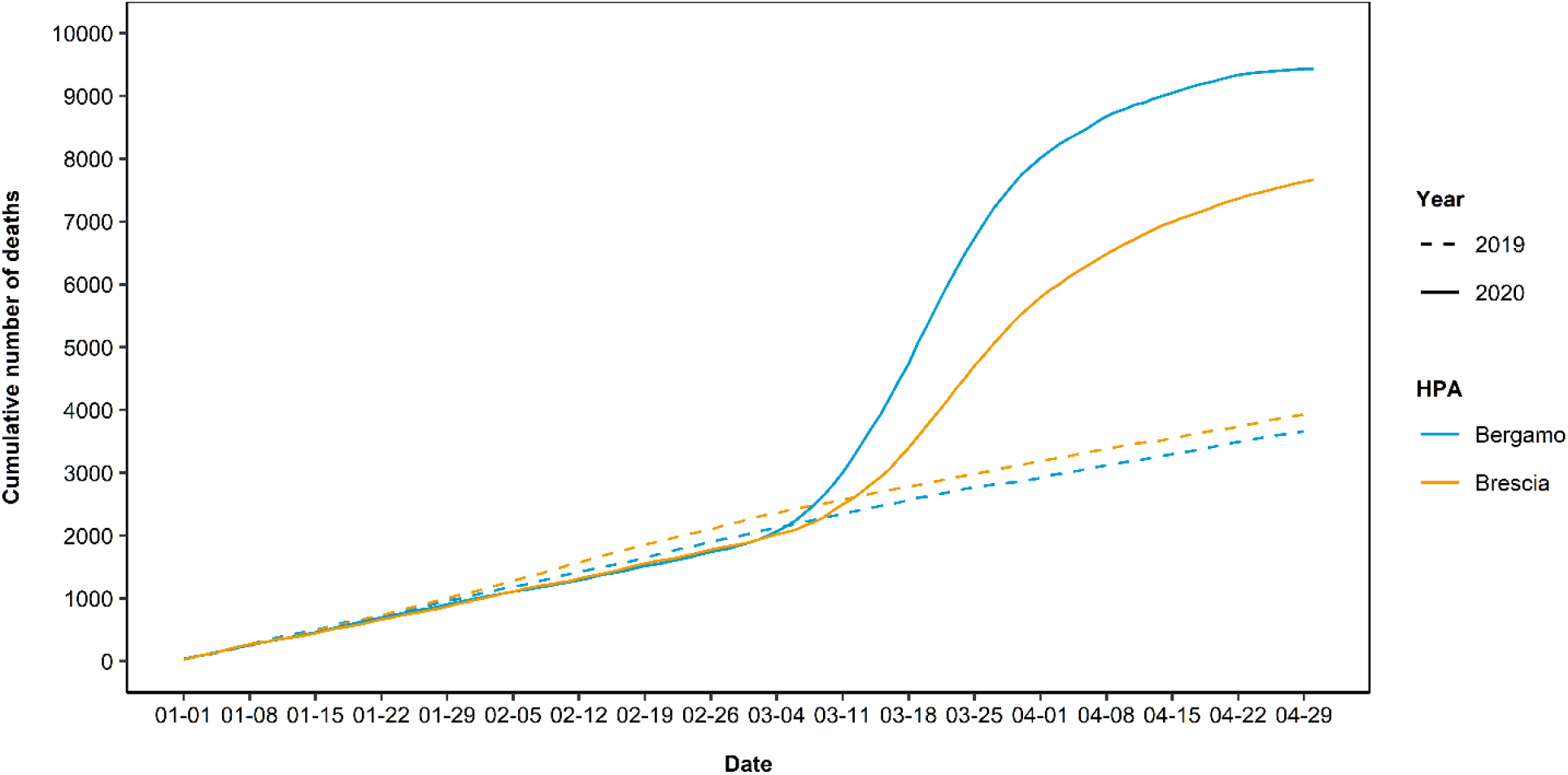
Comparison of 2020 and 2019 cumulative daily counts of death from January 1^st^ to April 30^th^, stratified by Health Protection Agency (HPA)

The trends in the estimated daily SMR confirm the substantial differences in the two HPAs for both sexes and for all the age classes older than 60 years (**Figure 2**). The overall daily estimated SMR peaked at 9.4 (95% CI: 9.1 – 9.7) in Bergamo province, and at 5.6 (95% CI: 5.4 – 5.8) in Brescia. The highest SMRs were observed among the elderly, peaking in the age-class 70-79: 12.7 (95% CI: 11.9 – 13.5) in Bergamo; 7.3 (95% CI: 6.8 – 7.9) in Brescia.

**Figure 2.**
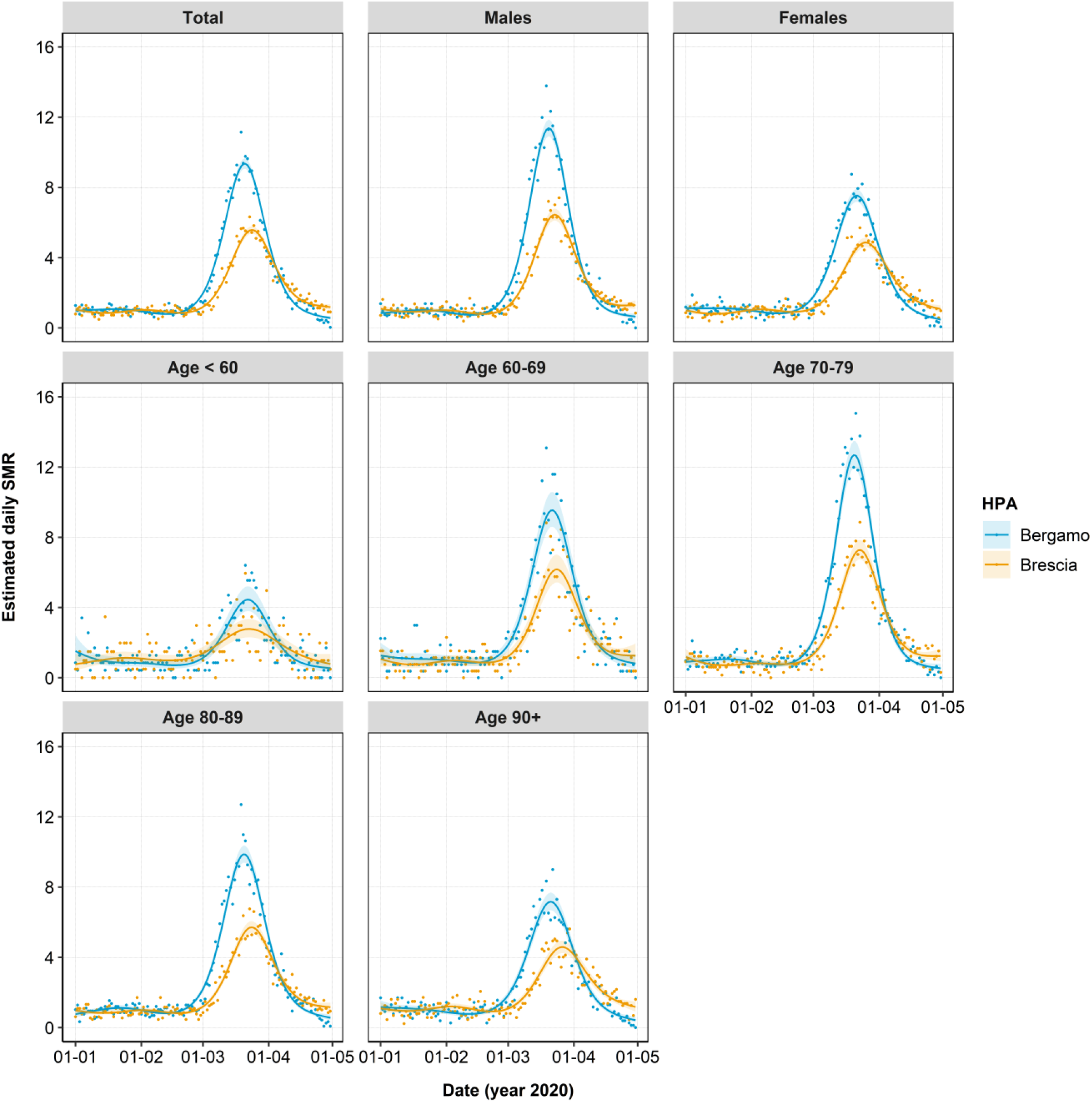
Predicted Standardized Mortality Ratio (SMR) from January 1^st^ to April 30^th^ 2020. Results from the overall analysis and from analyses stratified by age or sex, by Health Protection Agency (HPA)

Overall, during SARS-CoV-2 outbreak we estimated 5740 (95% CS: 5552–5936) excess deaths in the area of Bergamo, and 3703 (95% CS: 3535 – 3877) in Brescia, corresponding to a 2.55 (95% CS: 2.50 – 2.61) fold and 1.93 (95% CS: 1.89 – 1.98) fold increase in the number of cumulative deaths (**Table 2**; **Figure 3**).

**Table 2.** Estimated cumulative excess deaths and relative increase as compared to 2019, over the period January 1^st^ to April 30^th^ 2020, stratified by HPA, age or sex. HPA=Health Protection Agency. The summation of the estimated number of deaths within sex or age categories might not equal the total estimated excess, since stratified estimates are derived from separate models. CS=Credible Set.

| Disease | HPA Bergamo |  | HPA Brescia |  |
| --- | --- | --- | --- | --- |
|  | Estimated excess number of deaths (95% CS) | Estimated increase (n-fold) in the number of deaths (95%CS) | Estimated excess number of deaths (95% CS) | Estimated increase (n-fold) in the number of deaths (95%CS) |
| **Total** | 5740 (5552 - 5936) | 2.55 (2.50 - 2.61) | 3703 (3535 - 3877) | 1.93 (1.89 - 1.98) |
| **Sex** |  |  |  |  |
| Female | 2471 (2346 - 2601) | 2.28 (2.21 - 2.34) | 1724 (1609 - 1851) | 1.82 (1.77 - 1.88) |
| Male | 3269 (3134 - 3414) | 2.86 (2.78 - 2.94) | 1980 (1862 - 2103) | 2.06 (2.00 - 2.13) |
| **Age class** |  |  |  |  |
| <60 | 157 (120 - 203) | 1.55 (1.42 - 1.72) | 113 (80 - 155) | 1.46 (1.33 - 1.64) |
| 60-69 | 549 (496 - 612) | 2.70 (2.53 - 2.89) | 318 (274 - 374) | 2.01 (1.87 - 2.18) |
| 70-79 | 1520 (1429 - 1618) | 3.03 (2.91 - 3.17) | 920 (845 - 1009) | 2.16 (2.07 - 2.27) |
| 80-89 | 2398 (2283 - 2525) | 2.70 (2.62 - 2.79) | 1463 (1361 - 1577) | 1.94 (1.87 - 2.01) |
| 90+ | 1116 (1033 - 1212) | 2.20 (2.11 - 2.31) | 888 (807 - 981) | 1.84 (1.76 - 1.93) |

**Figure 3.**
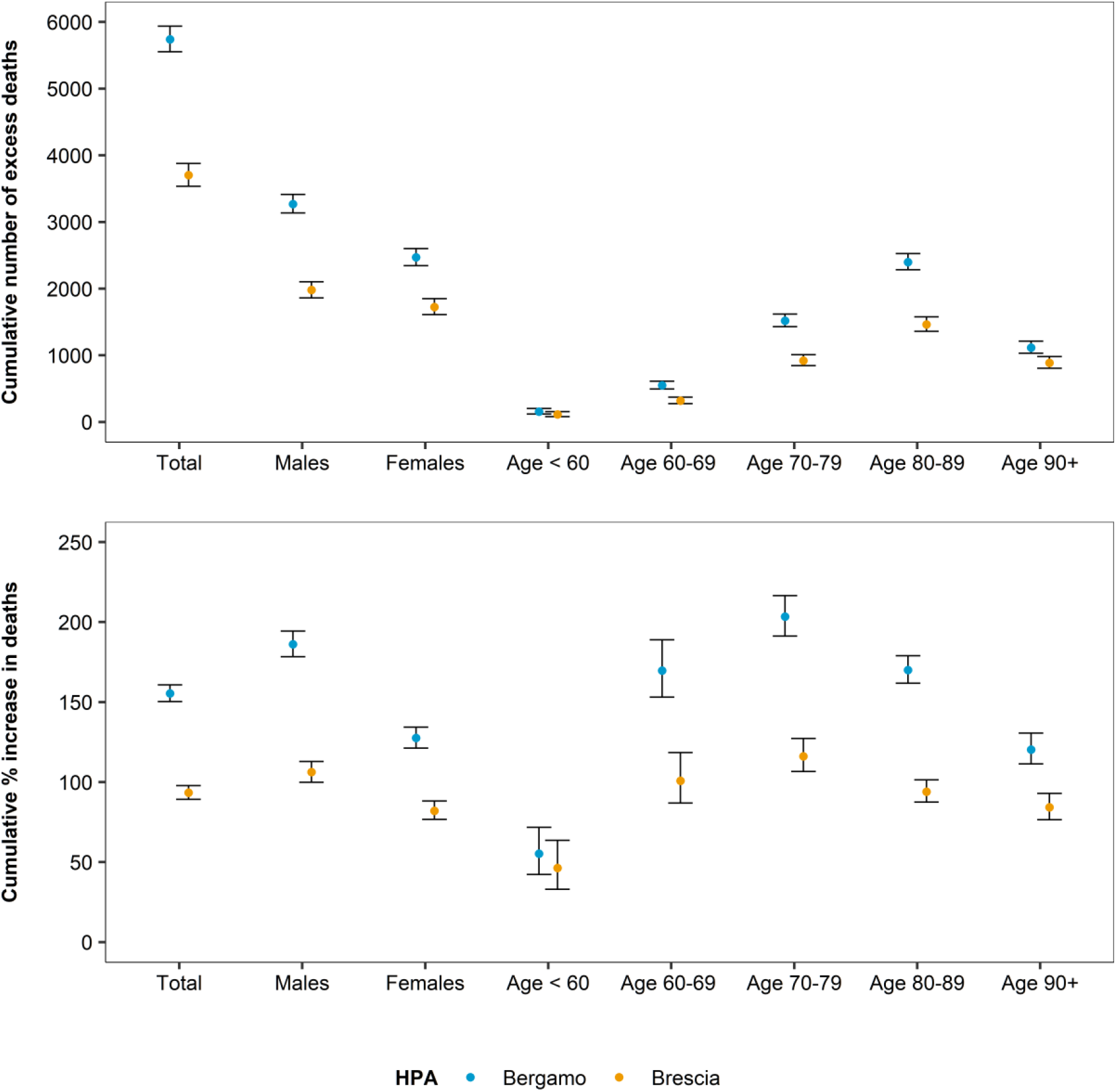
Estimated cumulative excess deaths and percent increase as compared to 2019, over the period January 1^st^ to April 30^th^ 2020, stratified by Health Protection Agency (HPA) and age or sex.

Restricting the analyses on the months of March and April, the estimated numbers of excess deaths were 5719 (95% CS: 5556 – 5892) in Bergamo and 3820 (95% CS: 3676 – 3967) in Brescia, representing virtually the whole excess mortality of the entire 2020 analyzed period (**Table 3**). This is confirmed by crude rates (**Table S1**): observed rates over the months of January and February during 2020 were comparable to those of 2019, if not slightly lower. The differential increase across sexes and age-classes was similar in the two provinces, but higher in Bergamo: in the age-class 70-79, the number of cumulative deaths increased by 5.05 folds (95% CS: 4.83 – 5.28), compared to 2.18 (95% CS: 1.96-2.44) among the youngest.

**Table 3.**
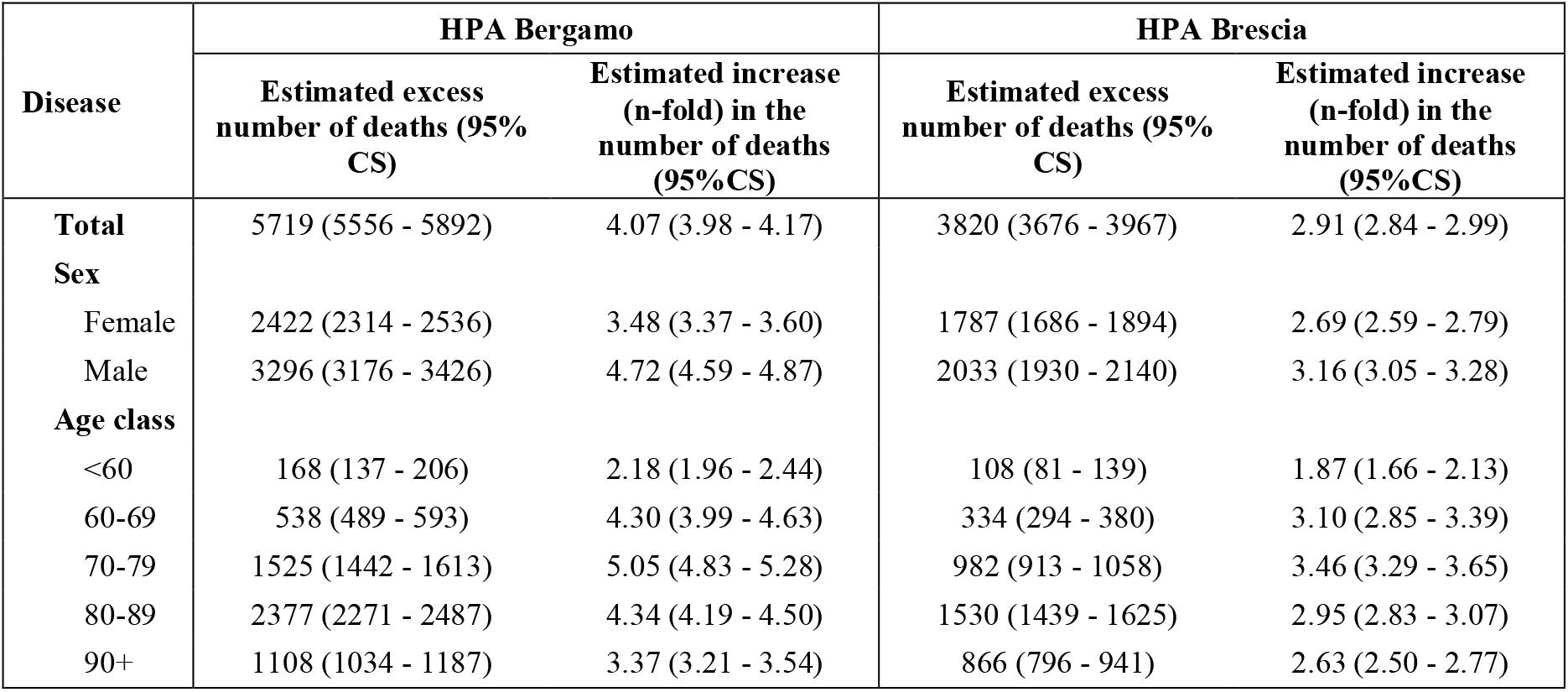
Estimated cumulative excess deaths and relative increase as compared to 2019, over the period March 1^st^ to April 30^th^ 2020, stratified by HPA, age or sex. HPA=Health Protection Agency. The sum of the estimated number of deaths within sex or age categories might not equal the total estimated excess, since stratified estimates are derived from separate models. CS=Credible Set.

Observing the time-series of the daily number of deaths (**Figure 4**), the excess death wave started earlier in the area of Bergamo, roughly one week after the detection of the first COVID-19 case in Lombardy (February 21^st^), and a few days later in the area of Brescia. In both areas, the excess death wave started before the introduction of lock-down measures (March 7^th^), and the peaks were observed between March 20^th^ and March 25^th^, roughly two weeks after the introduction of lock-down measures, returning to pre-epidemic levels by the end of April.

**Figure 4.**
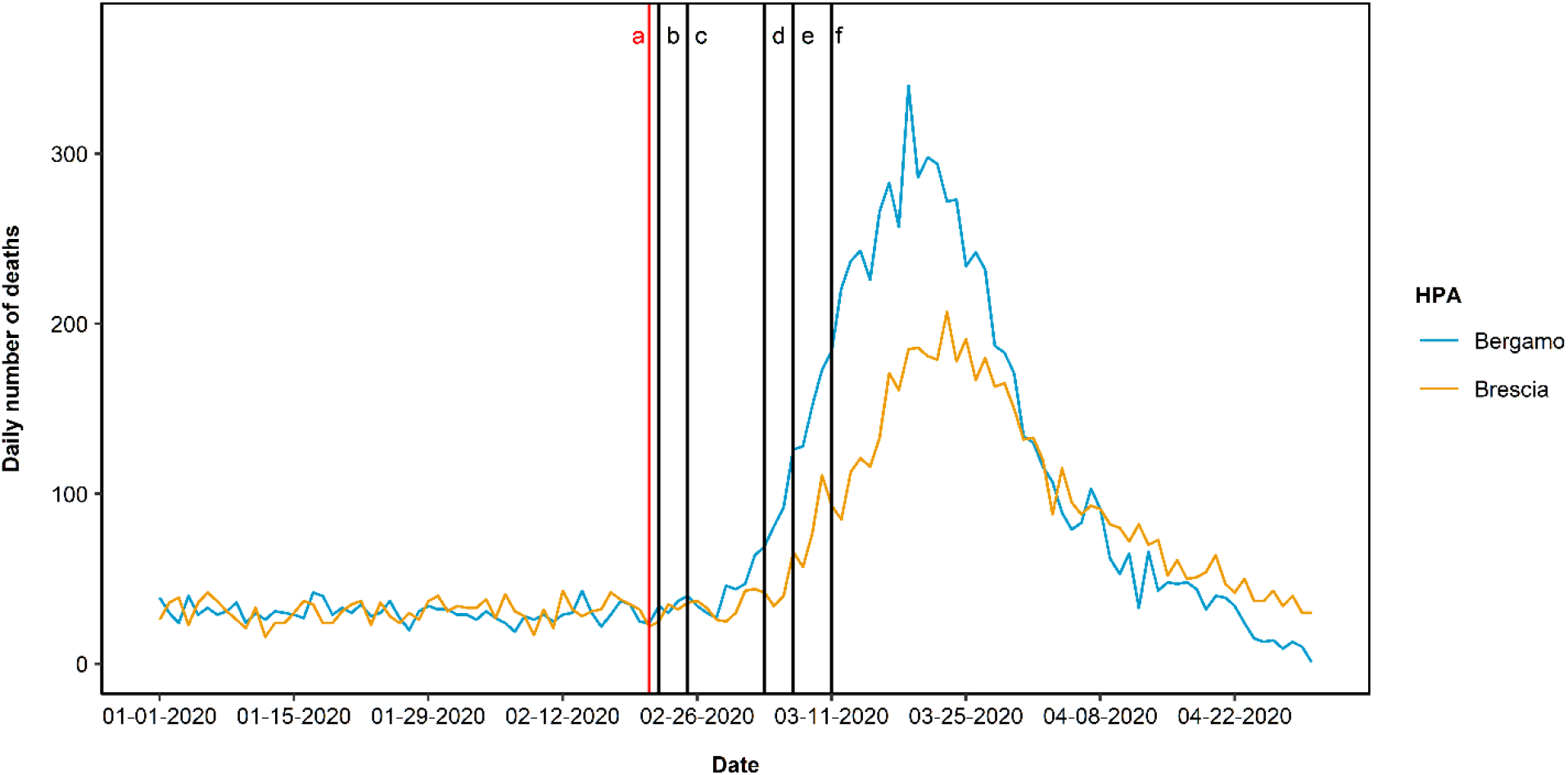
Time-series of daily number of deaths, stratified by Health Protection Agency (HPA). *a February 21*^*st*^: *first confirmed case in Lombardy* *b February 22*^*nd*^: *lock-down and quarantine for 11 municipalities* *c February 25*^*th*^: *school closed and crowd restrictions in 6 northern regions*, *d March 4*^*th*^: *school closed and crowd restrictions extended to the whole country e March 7*^*th*^: *lock-down and quarantine for Lombardy and 14 provinces* *f March 11*^*th*^: *lock-down and quarantine extended to the whole country*

## Discussion

This retrospective observational study, based upon the complete data of two Italian HPAs where the outbreak was particularly intense, evaluated the temporal trend of all-cause mortality during COVID-19 outbreak, describing its complete cycle using real world direct empirical observations.

During the entire 4-month period, in the areas of competence of the HPAs of Bergamo and Brescia, we estimated 9443 excess deaths, corresponding to a 2.2 folds increase as compared to those recorded in the same period of 2019, with a trend that rapidly rose starting 10 days after the first case of COVID-19 in the region. These results exceed by far the numbers of deaths formally attributed to the disease in the two HPAs, which registered respectively 2973 and 2295 deaths as of April 30^th^.

The large increase in the number of deaths could hardly be attributable to conditions other than COVID-19 epidemic, which affected mortality directly and potentially also indirectly, e.g. through, through the overcrowding of healthcare facilities and delayed care for time-dependent condition (such as stroke, myocardial infarction, etc.) [2,21]. Indeed, we observed that all excess deaths occurred in March and April, after the beginning of the disease outbreak, in line with preliminary findings considering only the month of March, which also suggested that the overall mortality did not increase in Italian provinces with low rates of infection [13].

Even though the two HPAs are adjacent, the wave of the daily number of deaths started a sharp increase a few days earlier in Bergamo than in Brescia; approximately two weeks before the introduction of the lock-down restrictions. These few days may have been crucial: although the waves had similar shapes, the province of Bergamo showed a significantly and substantially higher increase in mortality (2.55 folds), compared with Brescia (1.93 folds). These findings suggest that the national and local restrictions may have had a massive impact, resulting in a less steep wave in the time-series of the daily number of deaths in Brescia, where virus circulation was delayed. Moreover, in both HPAs we observed significant excess in mortality for eight weeks, with a peak four weeks after the start of the epidemic wave, and an overlapping descending phase. These trends indicated that the length of the excess death may not vary depending only on the circulation of the infection, which is correlated to overall SMR, suggesting that other factors might have influenced the overall mortality in both areas. One hypothesis can be identified in the emergency measures taken during the epidemic phase, like activation of Special Continuity Assistance Units (USCA), a primary care medical home service dedicated to COVID-19 patients, or the construction of field hospitals that lessened the overload of the healthcare facilities present in both areas [22,23].

Confirming previous findings, we observed an increased risk of death among males and the elderly [6-8,24]. Overall, the 70-79 years age group was the most affected in terms of cumulative deaths, although in COVID-19 patients the highest risk of death was observed among those aged 80 years or more [4]. The discrepancy might be due to a lower infection rate among the oldest, the mobility and social contacts of whom are frequently reduced. Noteworthy, we found a weaker excess mortality in nonagenarians than in younger elderly groups, possibly explained by the healthy survivor effect as well as a less clear-cut of direct and indirect COVID-19 impacts in those age-classes in which the underlying functional status is a more significant predictor of all-cause mortality than acute life-threatening conditions [25]. Moreover, demographics characteristics of the population may have contributed to design the curves, with the distribution of elderly Italian population that posed a challenge in reducing the impact of mortality due to the epidemic.

The strengths of the study include the use of administrative databases within a universal coverage system, on an unselected population of residents with complete knowledge on vital status, within an area where the outbreak has been particularly strong, and the virus circulated long before the enforcement of lock-down measures. Also, the analysis did not focus only on deaths classified as due to COVID-19, thereby avoiding underestimating the number of deaths at the beginning of the epidemic, when only a selected sample of patients underwent COVID-19 testing. Indeed, our estimates provide inferential estimates of excess deaths and SMRs, based on flexible models that could be easily reproducible in other geographical areas. Again, this is the first study describing the complete cycle of the outbreak: from the first cases to the regression to the baseline risk of death. This also allows the comparison of the excess death waves between two geographical areas, providing an initial description of how the timing of lock-down measures can ultimately impact on overall mortality. Finally, this research expressed robust data for evaluating the effectiveness of epidemic response and for providing informative measures to inform policy makers on the end of restrictive lockdown.

Some limitations should also be considered in interpreting the results. First, while accuracy is preserved, the precision of the estimated excess deaths is affected by the average number of daily events, decreasing in the subgroups that have low daily frequencies of death (e.g., the youngest age class). Second, the reference level is computed based on 2019 only: using a longer time-window could define the reference level better. However, trends in mortality within these areas proved to be very stable during the last years [26].

In conclusion, we documented a significant increase of the overall mortality during the first months of 2020, particularly March and April, indicating that COVID-19 outbreak had a substantially larger impact than what emerges from official estimates. Time-series analyses suggest that the national and local restrictions had a massive effect, determining a considerable reduction of COVID-19 burden. Furthermore, this study may serve as model for country-based estimations of overall (direct and indirect) impact of the COVID-19 epidemic on population mortality.

## Data Availability

NA

## Funding

None

## Conflict of interest

Nothing to disclose

## Supplementary material

**Table S1.** Crude rates over the period January 1^st^ to April 30^th^ 2020 in the two HPAs, overall and stratified by sex and/or age class. 95% Confidence Intervals were computed based on Poisson exact method.

|  | Crude rates (95% Confidence Intervals)*1000 person/months |  |  |  |  |  |
| --- | --- | --- | --- | --- | --- | --- |
|  | January-February | 2019<br>March-April | January-April | January-February | 2020<br>March-April | January-April |
| **HPA of Bergamo** |  |  |  |  |  |  |
| **Total** | 0.93 (0.89 - 0.98) | 0.78 (0.74 - 0.81) | 0.85 (0.83 - 0.88) | 0.85 (0.81 - 0.89) | 3.46 (3.38 - 3.54) | 2.17 (2.12 - 2.21) |
| **Sex** |  |  |  |  |  |  |
| Females | 0.99 (0.94 - 1.06) | 0.78 (0.73 - 0.84) | 0.89 (0.85 - 0.93) | 0.92 (0.86 - 0.98) | 3.07 (2.97 - 3.18) | 2.00 (1.95 - 2.06) |
| Males | 0.87 (0.82 - 0.93) | 0.77 (0.72 - 0.82) | 0.82 (0.78 - 0.86) | 0.79 (0.73 - 0.84) | 3.85 (3.73 - 3.97) | 2.33 (2.27 - 2.40) |
| **Age class** |  |  |  |  |  |  |
| <60 | 0.10 (0.08 - 0.11) | 0.08 (0.07 - 0.10) | 0.09 (0.08 - 0.10) | 0.08 (0.07 - 0.10) | 0.19 (0.17 - 0.22) | 0.14 (0.13 - 0.15) |
| 60-69 | 0.68 (0.58 - 0.79) | 0.58 (0.49 - 0.68) | 0.63 (0.56 - 0.70) | 0.66 (0.57 - 0.77) | 2.65 (2.46 - 2.85) | 1.66 (1.56 - 1.78) |
| 70-79 | 1.96 (1.76 - 2.17) | 1.88 (1.69 - 2.08) | 1.92 (1.78 - 2.06) | 1.81 (1.62 - 2.00) | 9.32 (8.90 - 9.74) | 5.59 (5.36 - 5.83) |
| 80-89 | 7.94 (7.39 - 8.53) | 6.46 (5.97 - 6.98) | 7.19 (6.82 - 7.58) | 6.51 (6.04 - 7.01) | 27.86 (26.89 - 28.86) | 17.27 (16.73 - 17.83) |
| 90+ | 34.18 (31.29 - 37.25) | 25.77 (23.31 - 28.41) | 29.90 (28.00 - 31.90) | 22.98 (20.94 - 25.17) | 76.75 (73.02 - 80.64) | 50.09 (47.94 - 52.31) |
| **Sex and age class** |  |  |  |  |  |  |
| **Females** |  |  |  |  |  |  |
| <60 | 0.08 (0.06 - 0.11) | 0.06 (0.04 - 0.08) | 0.07 (0.06 - 0.09) | 0.08 (0.06 - 0.10) | 0.12 (0.10 - 0.15) | 0.10 (0.08 - 0.12) |
| 60-69 | 0.53 (0.41 - 0.68) | 0.39 (0.29 - 0.51) | 0.46 (0.38 - 0.55) | 0.52 (0.41 - 0.66) | 1.21 (1.03 - 1.41) | 0.87 (0.76 - 0.99) |
| 70-79 | 1.44 (1.22 - 1.69) | 1.27 (1.07 - 1.50) | 1.35 (1.20 - 1.52) | 1.59 (1.36 - 1.85) | 5.45 (5.02 - 5.90) | 3.53 (3.29 - 3.79) |
| 80-89 | 6.65 (6.02 - 7.34) | 5.44 (4.87 - 6.05) | 6.04 (5.61 - 6.49) | 5.55 (5.00 - 6.14) | 20.96 (19.88 - 22.07) | 13.31 (12.71 - 13.94) |
| 90+ | 32.25 (29.08 - 35.66) | 23.86 (21.20 - 26.78) | 27.99 (25.90 - 30.19) | 21.46 (19.21 - 23.89) | 71.27 (67.15 - 75.56) | 46.57 (44.20 - 49.02) |
| **Males** |  |  |  |  |  |  |
| <60 | 0.11 (0.09 - 0.14) | 0.10 (0.08 - 0.13) | 0.11 (0.09 - 0.12) | 0.09 (0.07 - 0.11) | 0.26 (0.23 - 0.30) | 0.18 (0.16 - 0.20) |
| 60-69 | 0.83 (0.68 - 1.01) | 0.79 (0.64 - 0.96) | 0.81 (0.70 - 0.93) | 0.80 (0.66 - 0.97) | 4.14 (3.79 - 4.50) | 2.48 (2.30 - 2.68) |
| 70-79 | 2.56 (2.24 - 2.92) | 2.59 (2.27 - 2.94) | 2.58 (2.35 - 2.82) | 2.06 (1.78 - 2.37) | 13.78 (13.04 - 14.54) | 7.96 (7.57 - 8.38) |
| 80-89 | 10.13 (9.11 - 11.23) | 8.19 (7.29 - 9.17) | 9.14 (8.46 - 9.86) | 8.04 (7.21 - 8.95) | 38.81 (36.97 - 40.72) | 23.56 (22.54 - 24.61) |
| 90+ | 40.91 (34.37 - 48.33) | 32.40 (26.70 - 38.95) | 36.58 (32.19 - 41.40) | 27.93 (23.38 - 33.10) | 94.59 (86.12 - 103.66) | 61.53 (56.67 - 66.70) |
| **HPA of Brescia** |  |  |  |  |  |  |
| **Total** | 0.97 (0.93 - 1.02) | 0.75 (0.72 - 0.79) | 0.86 (0.83 - 0.89) | 0.81 (0.78 - 0.85) | 2.50 (2.43 - 2.56) | 1.66 (1.62 - 1.70) |
| **Sex** |  |  |  |  |  |  |
| Females | 1.02 (0.96 - 1.08) | 0.78 (0.73 - 0.83) | 0.90 (0.86 - 0.94) | 0.85 (0.80 - 0.91) | 2.40 (2.31 - 2.49) | 1.63 (1.58 - 1.69) |
| Males | 0.93 (0.87 - 0.99) | 0.72 (0.67 - 0.77) | 0.82 (0.79 - 0.86) | 0.77 (0.72 - 0.83) | 2.59 (2.50 - 2.69) | 1.69 (1.64 - 1.74) |
| **Age class** |  |  |  |  |  |  |
| <60 | 0.08 (0.07 - 0.10) | 0.06 (0.05 - 0.08) | 0.07 (0.06 - 0.08) | 0.08 (0.06 - 0.09) | 0.14 (0.12 - 0.16) | 0.11 (0.10 - 0.12) |
| 60-69 | 0.67 (0.58 - 0.78) | 0.52 (0.44 - 0.61) | 0.59 (0.53 - 0.66) | 0.53 (0.45 - 0.63) | 1.81 (1.66 - 1.98) | 1.18 (1.09 - 1.27) |
| 70-79 | 1.99 (1.81 - 2.19) | 1.64 (1.48 - 1.82) | 1.81 (1.69 - 1.95) | 1.55 (1.39 - 1.72) | 6.20 (5.88 - 6.54) | 3.89 (3.71 - 4.08) |
| 80-89 | 7.19 (6.72 - 7.69) | 5.47 (5.06 - 5.89) | 6.31 (6.00 - 6.64) | 5.53 (5.13 - 5.96) | 17.94 (17.21 - 18.68) | 11.79 (11.37 - 12.21) |
| 90+ | 21.99 (20.25 - 23.84) | 16.44 (14.96 - 18.02) | 19.17 (18.02 - 20.37) | 19.31 (17.73 - 20.99) | 47.86 (45.37 - 50.44) | 33.70 (32.22 - 35.23) |
| **Sex and age class** |  |  |  |  |  |  |
| **Females** |  |  |  |  |  |  |
| <60 | 0.07 (0.05 - 0.09) | 0.05 (0.04 - 0.07) | 0.06 (0.05 - 0.07) | 0.06 (0.04 - 0.08) | 0.09 (0.07 - 0.11) | 0.07 (0.06 - 0.09) |
| 60-69 | 0.39 (0.29 - 0.51) | 0.32 (0.23 - 0.43) | 0.35 (0.29 - 0.43) | 0.37 (0.28 - 0.49) | 0.99 (0.83 - 1.17) | 0.68 (0.59 - 0.79) |
| 70-79 | 1.58 (1.36 - 1.83) | 1.19 (1.00 - 1.40) | 1.38 (1.23 - 1.54) | 1.15 (0.96 - 1.36) | 3.90 (3.55 - 4.28) | 2.54 (2.34 - 2.75) |
| 80-89 | 6.05 (5.50 - 6.64) | 4.75 (4.27 - 5.26) | 5.39 (5.02 - 5.77) | 4.36 (3.91 - 4.85) | 14.44 (13.61 - 15.30) | 9.44 (8.97 - 9.93) |
| 90+ | 20.47 (18.57 - 22.52) | 15.46 (13.84 - 17.22) | 17.92 (16.67 - 19.25) | 19.05 (17.26 - 20.97) | 45.99 (43.21 - 48.90) | 32.63 (30.97 - 34.36) |
| **Males** |  |  |  |  |  |  |
| <60 | 0.09 (0.07 - 0.12) | 0.08 (0.06 - 0.10) | 0.09 (0.07 - 0.10) | 0.09 (0.07 - 0.11) | 0.19 (0.16 - 0.22) | 0.14 (0.12 - 0.16) |
| 60-69 | 0.97 (0.81 - 1.16) | 0.73 (0.59 - 0.89) | 0.85 (0.74 - 0.97) | 0.70 (0.56 - 0.86) | 2.68 (2.41 - 2.97) | 1.70 (1.54 - 1.86) |
| 70-79 | 2.47 (2.17 - 2.80) | 2.17 (1.89 - 2.48) | 2.32 (2.11 - 2.54) | 2.01 (1.74 - 2.30) | 8.84 (8.27 - 9.43) | 5.45 (5.13 - 5.78) |
| 80-89 | 9.04 (8.19 - 9.96) | 6.64 (5.92 - 7.41) | 7.82 (7.26 - 8.41) | 7.39 (6.65 - 8.19) | 23.45 (22.13 - 24.83) | 15.49 (14.72 - 16.28) |
| 90+ | 27.12 (23.15 - 31.58) | 19.75 (16.44 - 23.54) | 23.38 (20.77 - 26.22) | 20.14 (16.90 - 23.82) | 53.89 (48.54 - 59.67) | 37.15 (33.99 - 40.54) |

Stampa Messaggio

10/7/2020

**Inviato il:** 10-lug-2020 10.23

**Oggetto:** ERJ Open Research - Accept Decision on Manuscript ID ERJOR-00458-2020.R1

10-Jul-2020

Dear Dr. FERRARA:

It is a pleasure to accept your manuscript entitled “Magnitude and time-course of excess mortality during COVID-19 outbreak: population-based empirical evidence from highly impacted provinces in northern Italy” in its current form for publication in ERJ Open Research.

The comments of the reviewer(s) who reviewed your manuscript are included at the foot of this letter.

Some information on data sharing:

The ERS research journals encourage data sharing through the Dryad data repository (http://datadryad.org/). If you wish to deposit your dataset with Dryad, please ensure you specify that the dataset is linked to the ERJ Open Research and your manuscript ID: ERJOR-00458-2020.R1. The ERS has allocated funds to cover deposit fees, provided Dryad can link your submission to your article. For more information on data sharing please view our website (https://www.ersjournals.com/authors/data-sharing).

Thank you for your fine contribution. On behalf of the Editors of ERJ Open Research, we look forward to your continued contributions to the Journal.

Sincerely,

Prof. Alyn Morice

Chief Editor - ERJ Open Research

Associate Professor Woo-Jung Song

Deputy Chief Editor - ERJ Open Research

Dr. Alan Rigby

Associate Editor - ERJ Open Research

Editor Comments to Author:

Associate Editor

Comments to the Author:

Nothing further to add.

Reviewer(s)’Comments to Author:

## Notes

### Competing Interest Statement

The authors have declared no competing interest.

